# Longitudinal remote sleep and cognitive research in older adults with mild cognitive impairment and dementia: a prospective feasibility cohort study

**DOI:** 10.1101/2025.02.17.25322393

**Authors:** Victoria Grace Gabb, Jonathan Blackman, Hamish Duncan Morrison, Haoxuan Li, Adrian Kendrick, Nicholas Turner, Rosemary Greenwood, Bijetri Biswas, Elizabeth Coulthard

## Abstract

**INTRODUCTION:** We investigated the feasibility of remote longitudinal research using wearable devices and smartphone applications to record sleep and cognition in older adults with mild cognitive impairment (MCI) or dementia.

**METHODS:** Older adults with MCI or dementia due to Alzheimer’s disease (AD) or Lewy body disease (LBD) and cognitively healthy participants completed at-home sleep and circadian monitoring (digital sleep diaries, actigraphy, wearable sleep electroencephalography (EEG), saliva samples) and digital cognitive assessments for 8 weeks. Feasibility outcomes included recruitment, retention, and data completeness.

**RESULTS:** 41 participants consented (10 AD, 11 LBD, and 20 controls) and 40 completed the 8-week study. Data completeness for sleep EEG was 91% and ranged from 79% to 97% for all remote tasks. 12/40 (30%) participants reported receiving external support with completing study tasks.

**DISCUSSION:** Longitudinal multimodal sleep and cognitive profiling using novel technology is feasible in older adults with MCI and dementia and healthy older adults.

## 1 BACKGROUND

Sleep disturbances such as insomnia, fragmented sleep, daytime sleepiness, and sleep-disordered breathing are common features of Alzheimer’s disease (AD) and Lewy body disease (LBD) and often appear early in the disease course^1,2^. Short and fragmented sleep is associated with processes linked to neurodegeneration including glymphatic clearance of waste^3^, amyloid beta (Aβ) and tau deposition^4^, neuroinflammation^5^, and impaired cardiovascular health^6^, as well as neuropsychological and cognitive outcomes^7,8^. Several small studies of sleep apnoea treatment have demonstrated improvement in cognition and blood biomarkers of amyloid and tau burden, suggesting that sleep interventions could improve prognosis in MCI and dementia due to AD^9-11^. Poor quality or insufficient sleep is also increasingly considered a potentially modifiable risk factor for all-cause dementia and mild cognitive impairment (MCI)^12,13^, opening up the possibility that improving sleep in mid-life may protect against dementia. Large-scale studies are needed to confirm the most promising targets for sleep intervention and determine which treatments are most effective^14,15^.

Selecting sleep assessment tools is challenging, particularly in a population with cognitive impairment. Self-report is convenient, inexpensive, and scalable, and has often been used to examine sleep in people with dementia and MCI^16^. However, self-report correlates poorly with objective sleep measures, particularly in participants with MCI or dementia^17,18^ and those with subjectively poor sleep^19^, and cannot inform on key components of sleep such as sleep staging or micro-architecture. Polysomnography (PSG) is typically considered the gold standard for sleep measurement, as it provides rich objective sleep data. However, PSG typically requires expert set-up, analysis, and a controlled clinic environment, meaning it is expensive, not easily scalable, and therefore typically used for one or a few nights^20,21^. PSG set-up, especially in an artificial environment, may also not reflect natural sleep^21^.

Longitudinal data collection in sleep research would be beneficial for monitoring clinical trials and disease progression, and is also important to account for variation in sleep from external factors, such as acute illness or stress, as well as naturalistic intra-individual sleep variation^21^.

Wearable devices, smartphone applications and telemedicine, with the exception of actigraphy devices, have rarely been utilised in MCI or dementia research^22^, but offer an opportunity to collect objective and subjective data longitudinally in a natural setting^23^, often at relatively low-cost. Detailed and accurate sleep analysis can now be achieved through wireless technology, including electroencephalography (EEG) headbands and overnight pulse oximetry^23,24^. While actigraphy has been used in dementia research, EEG headbands and pulse oximetry are less well tested, especially in the earlier stages of impairment^25-27^. There is also increasing interest in remote cognitive testing and digital biomarkers for diagnosis and monitoring progression^28^. Despite this, the adoption of digital health technologies into neurology clinical trials, particularly in older people, has been slow^29^. Studies which have tested wearable devices and digital health technologies for sleep and dementia research in the home have typically collected feasibility data for a single device across only a few nights^30^ and required support from a study partner or care home staff^31-33^.

Remote sleep and cognitive data collection has potential to de-centralise clinical trials, making research more convenient for participants and reducing the costs, participant burden, and carbon footprint associated with study visits, whilst enabling real-time longitudinal data collection of treatment effects. Improved digital access and literacy among older adults^34^ and the use of technology among patients living with MCI/dementia to support independent living and for recreation indicates increasing acceptance of technology^35^. However, not all older adults are comfortable using technology, and changes to cognition, sensory processing, and communication might negatively impact the usability and acceptability of novel devices for research purposes in people with cognitive impairment^35,36^. Before trials invest in digital health technologies and remote study designs, it is important to know whether participants can and are willing to engage in such studies, and how much support might be required.

This study aimed to establish feasibility of predominantly technology-based sleep and cognitive assessments longitudinally from home in older adults with and without MCI/dementia.

## 2 METHODS

The Remote Evaluation of Sleep To enhance understanding of Early Dementia (RESTED) study was a prospective observational cohort study which recruited community-dwelling participants with MCI or dementia due to probable AD or LBD and age-matched cognitively healthy controls. Participants were asked to complete various sleep and cognitive tasks from home over an eight-week period.

The full study protocol has been published online^37^. The study has been reported in line with the Strengthening the Reporting of Observational Studies in Epidemiology (STROBE) guidelines (Supplementary Materials 1) and was approved by the Health Research Authority (Yorkshire and the Humber—Bradford Leeds Research Ethics Committee, reference 21/YH/0177). This study was conducted in accordance with the principles of the Declaration of Helsinki. All participants provided written informed consent prior to engaging in any study activities.

### 2.1 Study population

The RESTED study recruited community-dwelling adults aged 50 years or older. All participants were required to have internet access at home and not considered to have advanced dementia which would preclude them from completing the study tasks. Participants were recruited to one of three participant subgroups according to clinical diagnosis meeting standardised diagnostic criteria^38-41^. Participants with MCI or mild dementia due to probable AD were recruited to the AD group, participants with MCI or mild dementia due to probable LBD (including dementia with Lewy bodies and Parkinson’s disease (PD) MCI or dementia) were recruited to the LBD group, and sex- and age-matched individuals with no known neurodegenerative conditions or cognitive impairment were recruited as controls. Exclusion criteria included being in receipt of end-of-life care, acute illness, and significant comorbidities which might interfere with sleep, except for sleep disorders related to MCI/dementia or individuals undergoing treatment for OSA. There were no other restrictions on comorbidities.

Participants were recruited from the city of Bristol in United Kingdom and the surrounding areas via cognitive and movement disorders clinics and research volunteer databases, including Join Dementia Research. Our original target sample size of 75 participants^42^. However, due to the coronavirus pandemic, the study was delayed, and the budget was partially re-allocated to studies which required no patient contact^15,43,44^. Therefore, we revised the target sample size to 40 participants and recruited from a single site (North Bristol NHS Trust).

We did not recruit, nor require participants to have, a study partner. Participants were welcome to invite someone to support them during study appointments and throughout the study, and study support was recorded.

### 2.2 Study procedures

#### 2.2.1 Screening and baseline

Participants were pre-screened for eligibility over the telephone by a member of the research team. Prospective participants were invited to complete consent and screening in-person. Participants who scored less than 11/30 on the Montreal Cognitive Assessment (MoCA) at screening were considered too clinically impaired to participate in the study procedures and were withdrawn. Eligible participants completed baseline questionnaires with a researcher to assess sleep quality (Pittsburgh Sleep Quality Inventory, PSQI^45^), daytime sleepiness (Epworth Sleepiness Scale, ESS^46^), OSA risk (STOP-Bang^47^), symptoms of depression (Geriatric Depression Scale – 15 item, GDS^48^), anxiety (Generalised Anxiety Disorder - 7 item, GAD-7^49^), and apathy (Apathy Evaluation Scale – Self, AES-S^50^). Demographic information and medical histories were also recorded.

Participants were provided with a study kit (**Figure 1**) consisting of an actigraphy watch, wireless sleep EEG headband^25^, a USB charger, a home saliva collection kit for passive drool (for the dim-light melatonin assay) and oral swabs (for cortisol awakening response assay), and an overnight pulse oximeter. Participants who requested a study device were provided with a tablet for the duration of the study. Details on devices and the study kit are provided in **Supplementary Materials 2.** All participants were provided with a printed participant guide with instructions for each remote study task and research team contact details.

**Figure 1.**
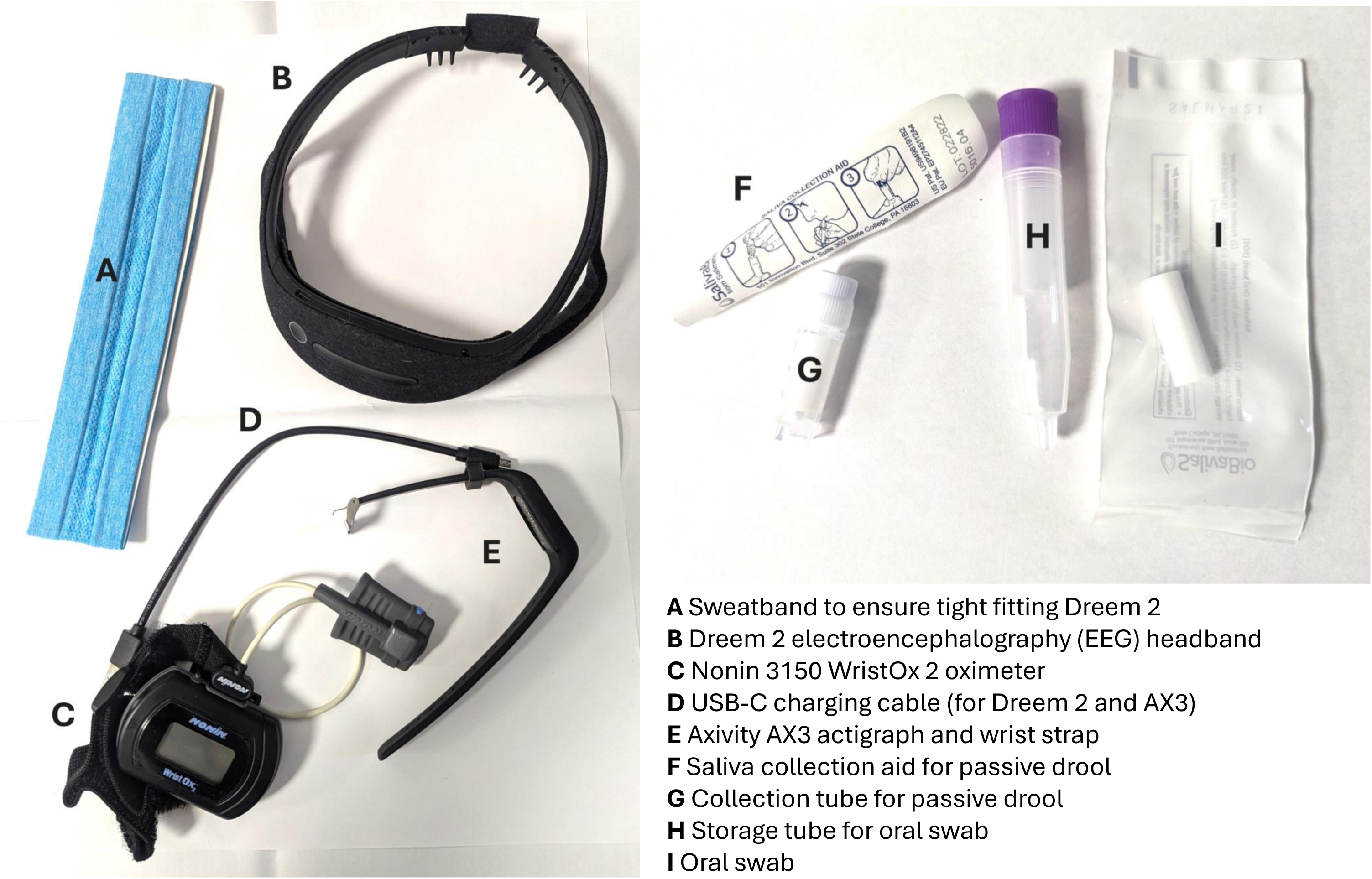
The study kit provided to RESTED participants. Participants were additionally required to use a smartphone or tablet to complete digital assessments.

At baseline, participants were provided with support to download and use the MyDignio application, a telemedicine cloud-based software which was used to deliver the digital sleep diaries, questionnaires during the main study period, and reminders to complete study tasks.

Participants were also familiarised with the study schedule and tasks, including the online cognitive tests, and offered support and training ad-hoc throughout the study.

#### 2.2.1 Main study period

Following baseline, participants completed 56 days of continuous sleep and regular cognitive monitoring in their own home. Participants were asked to wear an Axivity AX3 actigraphy watch, preferably on their non-dominant hand, and complete daily sleep diaries via the MyDignio application for 56 consecutive days. Sleep diaries consisted of the nine main Consensus Sleep Diary^51^ questions, with an additional question to gauge subjective sleep quality compared to their typical sleep: “Would you consider your sleep last night to have been much worse than normal?”.

Across the eight-week period, participants were scheduled to complete a set of three online cognitive tests twice weekly on an online assessment platform (Cognitron). The three cognitive tasks (choice reaction time, forward digit span, self-ordered search) were hosted on a bespoke RESTED study version of the Cognitron platform (http://www.cognitron.co.uk/).

For 7 days during this period, participants also completed an “intensive week”, consisting of daily cognitive tasks on Cognitron and nightly sleep recording using the Dreem 2, a wireless headband which records and stores EEG utilising three frontal (F7, F8, Fp1) and two occipital (O2, O1) dry electrodes. The Dreem sleep staging algorithm has comparable accuracy to manual sleep expert scoring of PSG^25^, including in cognitively healthy older adults as well as patients with AD^52^ and PD^53^. Sleep recordings were initiated by the participant at their natural bedtime each night and terminated after natural awakening each morning. Data was uploaded via Bluetooth and Wi-Fi to a server accessible to the research team.

During the same week, participants were asked to complete two memory assessments with a researcher via videoconferencing software. Each assessment consisted of two phases, one during the evening and one the following morning after sleep. During the evening phase, participants were given an encoding task during a word list (categorising words as “alive” or “not alive”), asked to free recall, then repeat the encoding task. On one of the evenings, participants also completed a separate autobiographical memory task (listing five things they did that day). During the morning phase, participants were tested on free recall and whether they could recognise the words they saw amongst a mixed list of target and distractor words. To assess the feasibility of remote data collection of biomarkers for circadian analysis, participants were asked to complete serial saliva samples across one evening (to assess dim-light melatonin onset) and one morning (to assess cortisol awakening response).

During the study, participants were also invited to undergo two nights of pulse oximetry for sleep apnoea screening and a blood test for plasma biomarker analysis of Aβ42:40, phosphorylated tau (p-tau) 181 and 217, neurofilament light chain (NFL), and glial fibrillary acidic protein (GFAP). A protocol amendment approved partway through the study introduced bespoke questionnaires probing study expectations, reasons for participation, experience with technology, and how acceptable they found the intensive week study tasks.

Participants were also asked to attend a remote end-of-study interview to share their experiences and asked to return for a 6-month follow-up to complete a MoCA and any outstanding study tasks (e.g., missed blood test). Findings from the interviews will be reported in detail elsewhere.

### 2.3 Feasibility and acceptability outcomes

#### 2.3.1 Recruitment and retention

Consent, recruitment, and retention rates were recorded and are summarised in a flowchart, alongside reasons for non-participation at each stage of the recruitment process.

#### 2.3.2 Data quality and completeness

For each remote study task, data completeness was assessed by the average number and percentage of completed tasks or nights’ use per participant per participant group (data completeness rate). The number and percentage of individuals who completed the maximum number of data points for a given study task (e.g., completed all requested 7 nights of EEG) is also provided.

For EEG, signal quality is reported based on the automated algorithm provided by Dreem, where optimal signal quality is considered ≥ 85% and good quality is considered ≥ 70%.

#### 2.3.3 Associations between participant characteristics and adherence

To explore potential associations between key clinical and demographic variables and adherence, we examined the correlation between two continuous measures of adherence (number of sleep diaries completed and mean record quality of Dreem EEG data) and four continuous variables which might impact digital literacy and/or engagement (age at consent, apathy, subjective sleep quality as assessed by the PSQI total score, and baseline cognitive impairment as assessed by MoCA total score).

#### 2.3.4 Study support and resource use

Support from outside the study team was quantified by the number of participants who attended study visits with someone to support, informed the study team that they had someone external to the study team who could support study tasks, and the number of participants who reported receiving support on at least one study task. The type of support from others and support provided by the research team is summarised descriptively.

### 2.4 Data analysis

Unless otherwise stated, descriptive statistics are provided as mean (standard deviation, SD) for continuous variables, and frequency and percentage for categorical variables, and provided for the full cohort as well as by participant subgroup (AD vs LBD vs controls). Data analysis was performed using R (version 4.3.1) and R Studio software (version 2023.6.0.421). Actigraphy data was processed using the open source AX3/AX3 OMGUI application and analysed in R using the *GGIR* package^54^. Where available, sleep timing was adjusted using participants’ sleep diaries, otherwise *GGIR* uses the Heuristic algorithm looking at Distribution of Change in Z-Angle (HDCZA)^55^ to estimate sleep timing. All files were also visually inspected to check for accuracy of sleep timing. Quality of the Dreem 2 recordings was assessed by inspecting Dreem’s automated record quality index in the sleep report for each night, which indicates the percentage of the recording that is of scorable quality for sleep analysis.

Associations between participant characteristics and adherence were calculated using Spearman’s rank correlation with alpha set to *p < 0.05*. Reasons for non-participation and missing data are provided where known and mapped to the Capability Opportunity Motivation model of behaviour change (COM-B model)^56^. Where additional data was collected on any outcome (e.g., participants completed an additional night of EEG than instructed), additional datapoints are removed prior to analysis to avoid biasing feasibility metrics.

### 2.5 Patient and public involvement

Patient and public involvement (PPI) was sought from individuals with lived experience of MCI and dementia before and throughout the study. PPI contributors reviewed and improved study documents and advised on the acceptability of adding blood biomarker testing. The PPI group strongly endorsed our additional recruitment materials (including an advertisement poster and a Plain English participant information sheet) that we introduced partway through the study after we received feedback from a prospective participant that standard participant information sheets are too long and use inaccessible language for people with cognitive impairment. We also introduced a “reduced” study protocol which involved use of a paper (rather than digital) sleep diary, actigraphy, and EEG, to encourage recruitment of participants who may be more comfortable with less frequent use of technology, however nobody chose the reduced protocol; they either declined altogether or participated in the full study.

## 3 RESULTS

### 3.1 Participant characteristics and enrolment

#### 3.1.1 Recruitment

Recruitment was open for 17 months from February 2022 to July 2023 with an average recruitment rate of 3 participants per month. Of 129 individuals identified as potentially eligible during pre-screening, 44 individuals consented to take part, giving a consent rate of 34% (**Figure 2**). Participants described different motivations for taking part, including wanting to support dementia research, perceiving the study as helpful for themselves and/or others, and because the study sounded interesting or novel. Reasons for declining to take part in the study are described in **Figure 3** and mapped to the Capability Opportunity Motivation-Behaviour (COM-B) model of behaviour change^56^.

**Figure 2.**
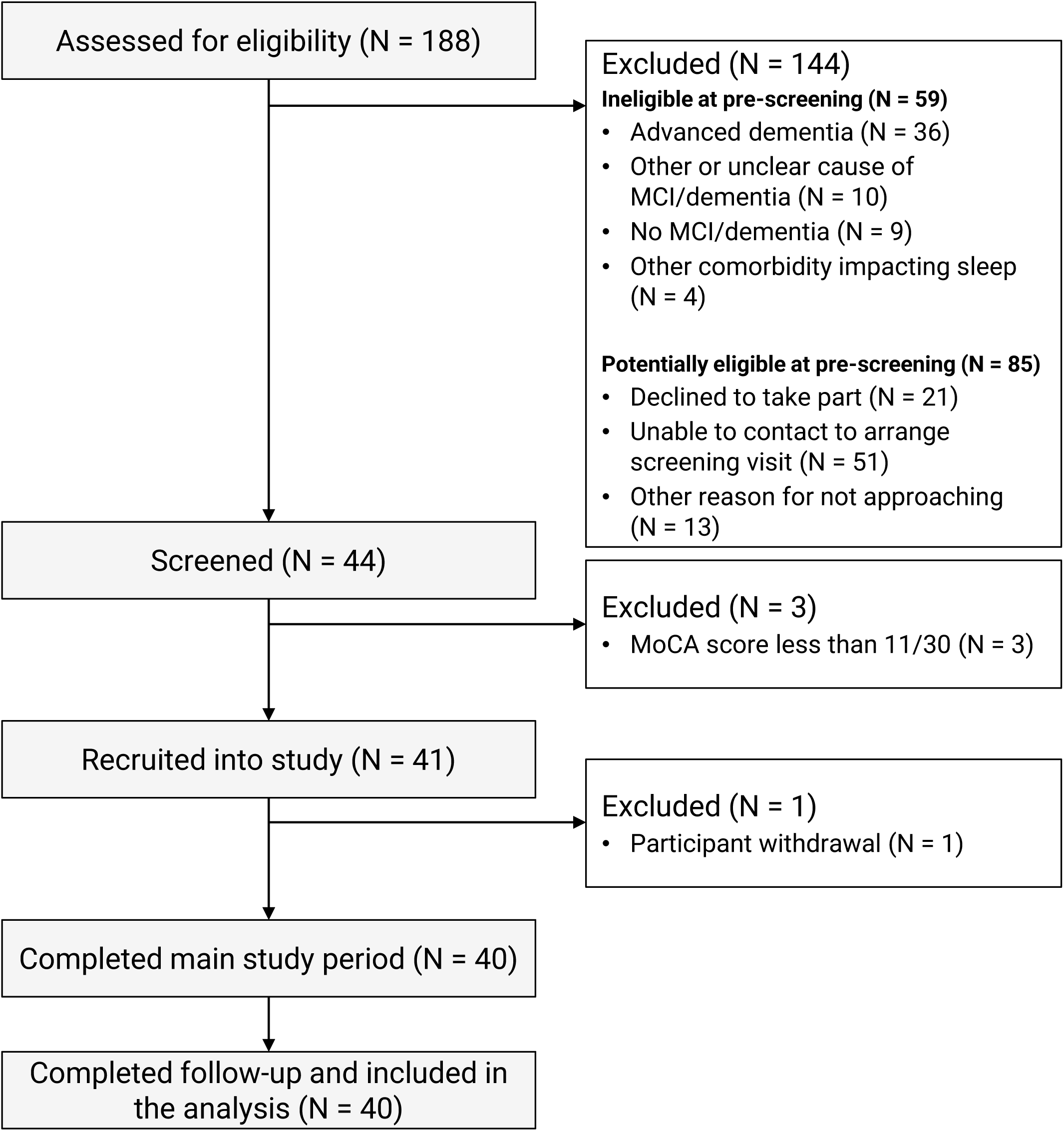
Participant flow through the RESTED study. Of 129 potentially eligible participants, 44 (34%) consented to participate and 41 (32%) were recruited. Retention was very high (98%) following recruitment to the study.

**Figure 3.**
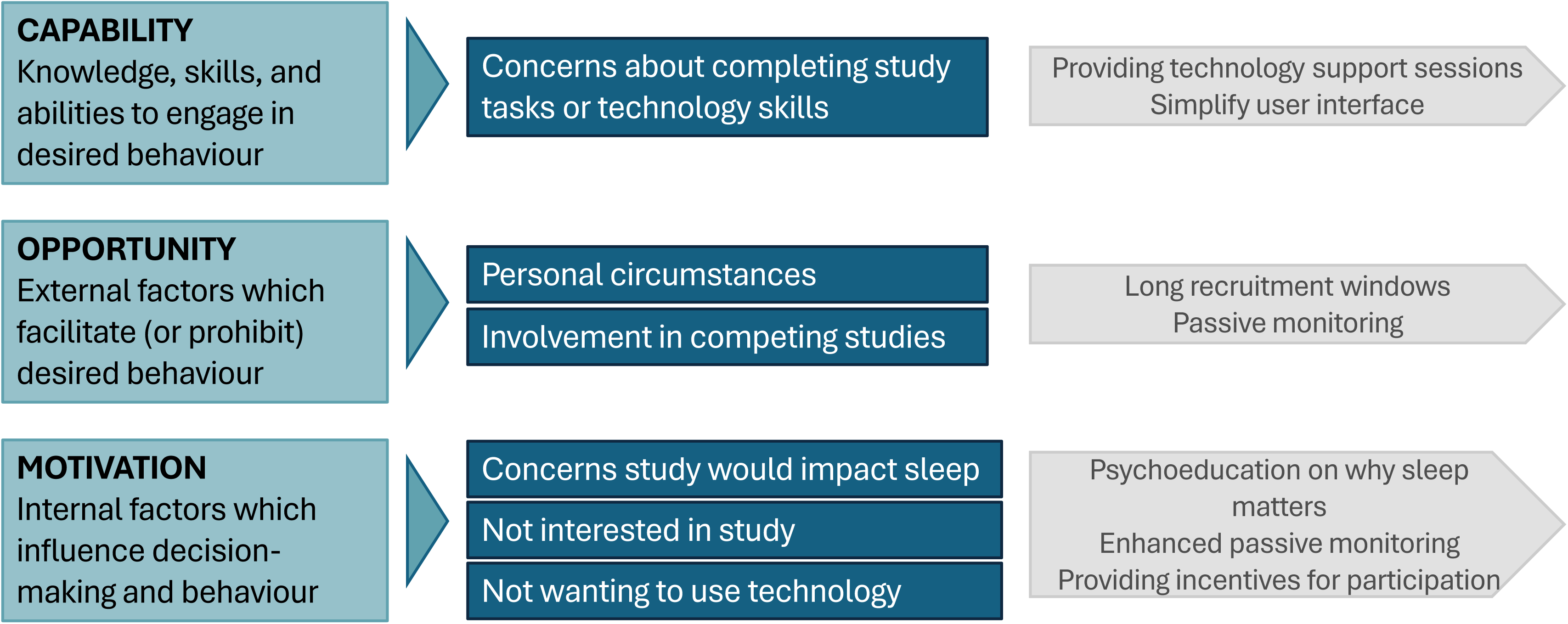
Reasons for declining to participate in the study could be categorised according to the Capability Opportunity Motivation-Behaviour (COM-B) model of behaviour change (blue). Opportunities to increase capability, opportunity, and motivation to take part in the research are outlined (grey).

#### 3.1.2 Retention

Of 44 individuals who completed screening, three scored less than 11 on the MoCA and were withdrawn. 41 participants were eligible and completed all baseline assessments and training in the at-home study tasks. One participant withdrew prior to starting the at-home study tasks due to personal circumstances and perceived study burden. All remaining 40 participants completed the main 8-week study period and 6-month follow-up and were included in the analysis, giving a retention rate of 98%.

#### 3.1.3 Participant characteristics

Participant demographics and baseline variables are presented in **Table 1**. The sample was predominantly male with a mean age at consent of 70.9 years (SD = 5.9, range = 57 to 81). Most participants were retired (82.5%) and all identified as White British. 25/40 participants completed the RESTED Expectations questionnaire, which was introduced via an amendment partway through the recruitment period. Of these, most participants reported frequent use of smartphones or other smart technology (18/25, 72%), however only 3/25 (12%) participants had previously used a wearable to monitor their sleep.

**Table 1.** Participant characteristics for the study cohort.

|  | AD group<br>N=10 | LBD group<br>N=10 | Control group<br>N=20 |
| --- | --- | --- | --- |
| **Diagnosis at consent<sup>1</sup>** |  |  |  |
| Mild cognitive impairment | 4 (40%) | 6 (60%) | 0 |
| Dementia | 6 (60%) | 4 (40%) | 0 |
| **Blood biomarkers of neurodegeneration/AD pathology<sup>1</sup>, pg/mL** |  |  |  |
| Aβ40:42 | 0.064 (0.006) | 0.062 (0.006) | 0.072 (0.007) |
| p-tau181 | 26.2 (7.1) | 24.9 (12.7) | 24.9 (12.7) |
| p-tau217 | 0.7 (0.4) | 0.8 (0.4) | 0.6 (0.4) |
| GFAP | 102.2 (28.1) | 140.9 (56.4) | 103.5 (48.1) |
| NFL | 23.9 (18.0) | 18.7 (6.5) | 13.4 (5.1) |
| **Age at consent, years** | 69.2 (7.9) | 73.9 (2.8) | 70.3 (5.7) |
| **Female sex<sup>1</sup>** | 2 (20%) | 2 (20%) | 5 (25%) |
| **Employment status<sup>1</sup>** |  |  |  |
| Full-time employment | 2 (20%) | 0 | 2 (10%) |
| Part-time employment | 0 | 0 | 3 (15%) |
| Retired | 8 (80%) | 10 (100%) | 15 (75%) |
| **Education<sup>1</sup>** |  |  |  |
| Secondary (≤ 12 years) | 4 (40%) | 5 (50%) | 4 (20%) |
| Further or higher (> 12 years) | 6 (60%) | 5 (50%) | 16 (80%) |
| **Co-morbid diagnoses<sup>1</sup>** |  |  |  |
| Obstructive sleep apnoea* | 0 | 1 (10%) | 2 (10%) |
| Musculoskeletal/pain | 3 (30%) | 3 (30%) | 8 (40%) |
| Urinary/prostate | 3 (30%) | 2 (20%) | 3 (15%) |
| Depression/anxiety | 1 (10%) | 0 | 1 (5%) |
| **Medications<sup>1</sup>** |  |  |  |
| Cognitive enhancer | 6 (60%) | 6 (60%) | 0 |
| Dopaminergic | 0 | 8 (80%) | 0 |
| Melatonin | 0 | 3 (30%) | 0 |
| Hypnotic/sedative | 0 | 2 (20%) | 1 (5%) |
| Antidepressant | 4 (40%) | 4 (40%) | 4 (20%) |
| **Baseline cognition<sup>2</sup>: MoCA** | 23.0 (4.9) | 22.0 (4.3) | 27.1 (1.4) |
| **Subjective sleep quality<sup>2</sup>: PSQI** | 4.3 (2.0) | 7.8 (3.0) | 6.0 (3.9) |
| **Self-reported anxiety<sup>2</sup>: GAD-7** | 5.6 (5.2) | 3.5 (2.1) | 3.6 (4.3) |
| **Self-reported depression<sup>2</sup>:**<br>GDS-15 | 4.5 (3.3) | 4.1 (2.3) | 2.2 (2.2) |
| **Self-reported apathy<sup>2</sup>: AES-S\*\*** | 62.2 (6.0) | 53.8 (9.9) | 62.8 (8.6) |

<sup>1</sup> Frequency (%); <sup>2</sup> Mean (SD); \*Participants with obstructive sleep apnoea were included only if they were on treatment with continuous positive airway pressure; \*\* One participant had a missing value for one item, so this score was imputed using simple imputation. *Abbreviations:* MoCA: Montreal Cognitive Assessment; PSQI: Pittsburgh Sleep Quality Index; GAD-7: Generalised Anxiety Disorder – 7; GDS-15: Geriatric Depression Scale – 15-item scale; AES-S: Apathy Evaluation Scale – Self; SD: standard deviation.

### 3.2 Data quality and completeness (adherence)

Adherence was very high across all three study groups (**Table 2**). Reasons for missing data are summarised in **Figure 4** with additional detail provided in **Supplementary Materials 3.**

**Figure 4.**
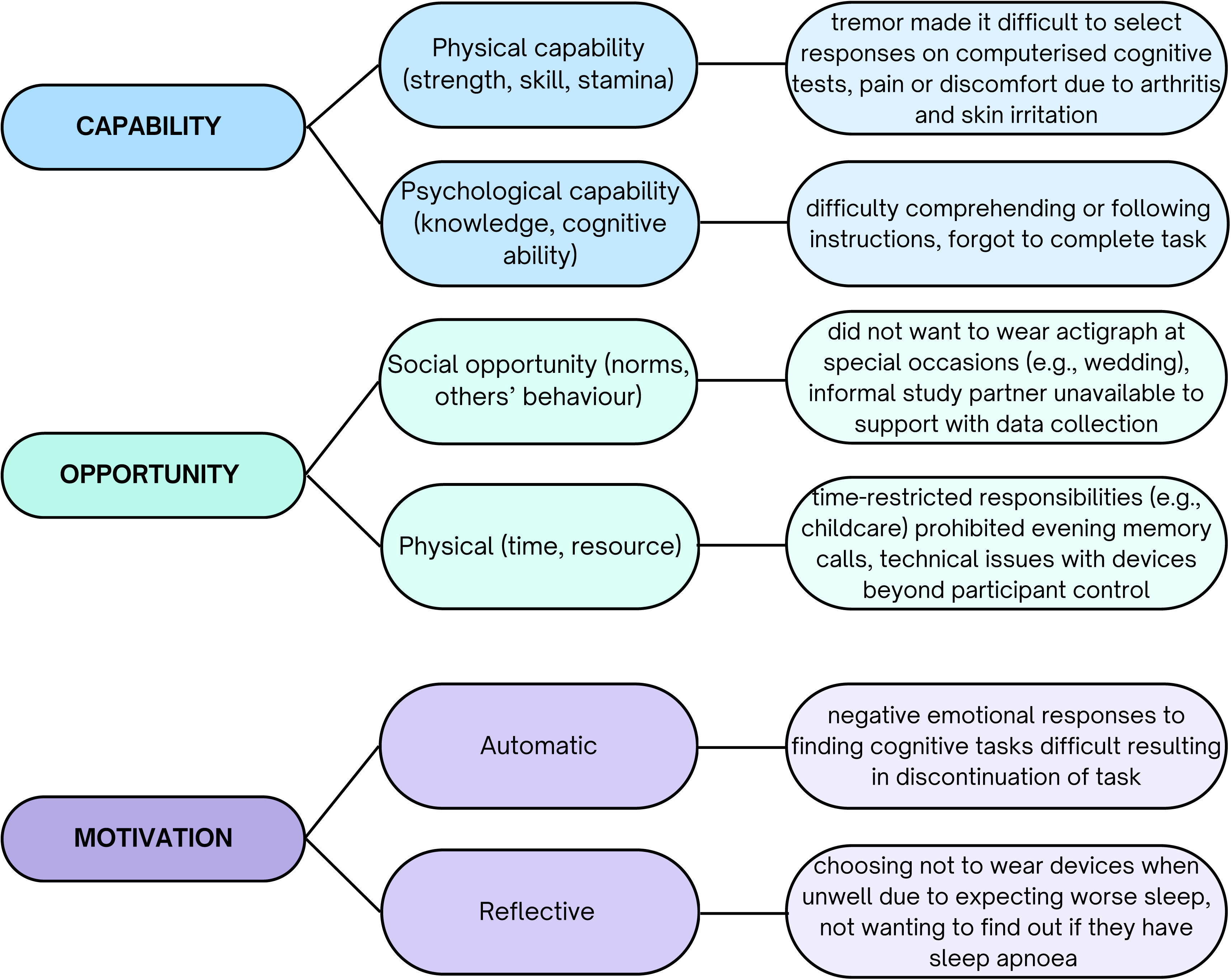
Reasons for missing data are mapped the Capability Opportunity Motivation-Behaviour (COM-B) model of behaviour change.

**Table 2.**
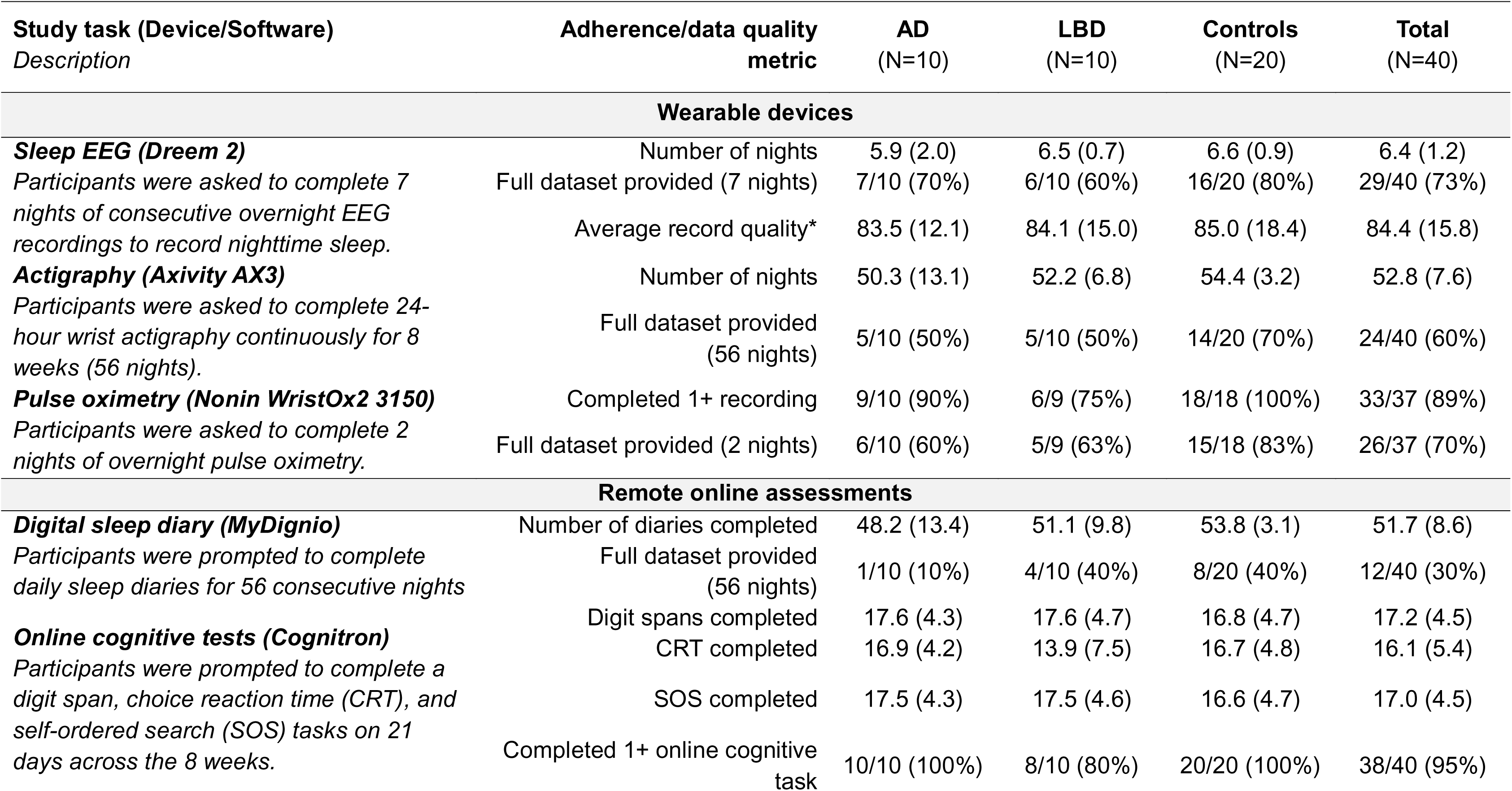
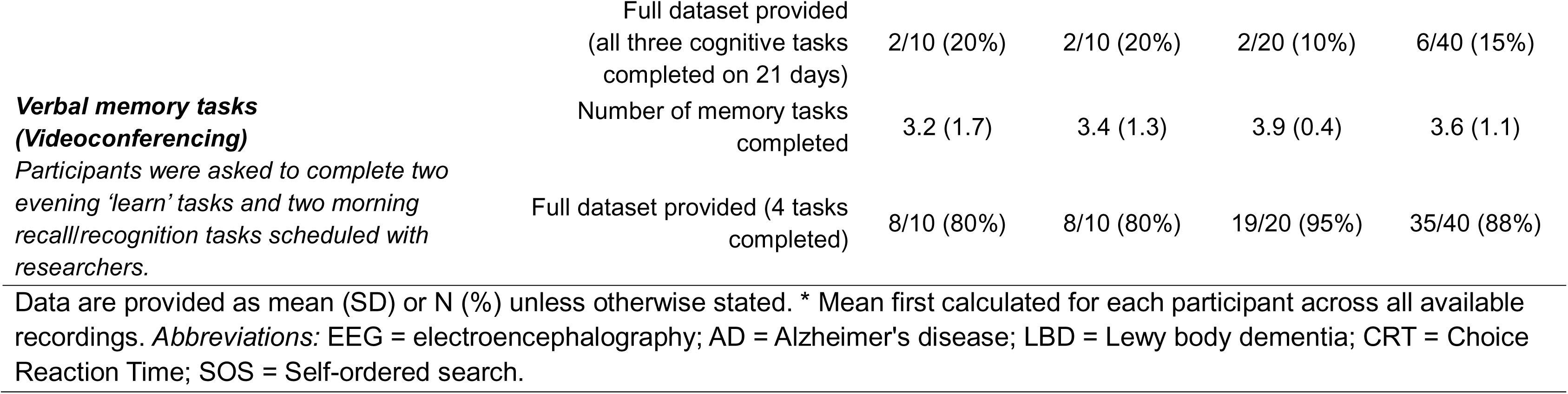
Summary of feasibility outcomes relating to data quality and adherence for wearable devices and remote/online cognitive assessments.

#### 3.2.1 Sleep EEG

In total, 257 recordings were made using the Dreem 2. One participant accidentally completed an eighth recording which was removed from subsequent analyses to reduce bias. All participants successfully recorded at least one night of sleep, with an average of 6.4 (1.2) nights across the cohort or 91.4% data completeness rate. 29/40 (73%) provided data for all 7 nights. Average record quality was also high at 84.4% (15.8), indicating nearly optimal record quality on average across the cohort. One participant wore the Dreem 2 but did not successfully initiate the recording during the intensive week but completed 7 successful nights of recording on a second attempt later in the study. Record quality for individual EEG channels is provided in **Supplementary Material 4**.

#### 3.2.2 Actigraphy

2,332 nights of data across the full cohort was collected. 189 of these nights contained no data, either at the beginning or the end of the actigraphy file and were removed from sleep analyses. Following visual inspection, a further 9 nights were removed due to sustained non-wear. Therefore, 2,134 nights were of sufficient quality for sleep analysis.

Across the cohort, participants provided an average of 52.8 (7.6) out of 56 analysable nights, giving a data completeness rate of 94.3%. 24/40 (60%) provided 56 analysable nights. All participants recorded at least 14 days’ of actigraphy. Unexpected battery failure outside of participants’ control (due to long-term storage without use during the coronavirus pandemic) affected two participants’ recordings and was identified during data check at the midpoint study visit. No participants refused to wear or prolonged discontinued use of the actigraphy device, though several reported forgetting or choosing not to wear it on certain days or taking brief breaks due to minor skin irritation.

37 diary entries were manually entered based on visual analysis of the actigraphy data due to missing sleep diaries and poor HDCZA algorithm detection of the sleep period. Finally, the sleep analysis software was instructed to solely rely on sleep diary information for 166 nights due to clear misclassification of sleep period.

#### 3.2.3 Overnight pulse oximetry

As three participants were already on treatment for established OSA, 37 participants were offered overnight pulse oximetry. Two participants (5.4%) declined, and one participant was unable to tolerate wearing the device. The remaining 33 (89.1%) participants completed at least one successful overnight recording, with 26 (70.3%) recorded successful oximetry traces on both nights. Data completeness rate was 79.7% for those eligible and asked to complete overnight pulse oximetry. Referrals to a sleep clinic for either sleep apnoea or incidental findings (such as abnormal pulse rise index) were indicated for 21/37 participants (56.8%). Four participants (10.8%) declined referral due to not wanting a formal diagnosis, not wanting to be put on sleep apnoea treatment, and inconvenience of travelling to the clinic. 17 (45.9%) participants agreed to a referral.

#### 3.2.4 Digital sleep diaries

Although few participants completed all 56 sleep diaries (12/40, 30%), overall adherence was high. Participants completed an average of 51.7 (8.6) sleep diaries and only two participants completed fewer than 75% of their sleep diaries. Data completeness rate was 92.3%.

One participant completed sleep diaries on paper whilst her partner who facilitated entries into MyDignio was away. Three participants had technical issues with data entry into the MyDignio app lasting several days due to a software update and provided some of their sleep diaries via email which were then entered by the study team.

At the time of study set-up, Dignio did not offer data format validation and free-text response boxes were used for several questions. Participants often did not utilise the format requested and inconsistency in reporting prohibited reliable automated re-coding, however most were manually interpretable by the research team. Where a range of values were provided, a mean was calculated and the value was rounded to the nearest whole minute (e.g., 5 to 10 minutes was re-coded to 8 minutes). One diary entry was manually corrected by the research team to align with other diary entries and the actigraphy file. It was not possible to re-code some responses due to ambiguity (e.g., responses of “not long”, “several minutes”, or “unsure”), leaving 43 completed sleep diaries incomplete on at least one sleep variable. Questions most likely to have a missing value were questions on sleep onset latency and length of nocturnal awakenings.

#### 3.2.5 Video-based verbal memory tasks

37/40 (92.5%) participants completed at least one completed verbal memory task with a researcher. Data was missing for one participant as the task was considered inappropriate due to their specific language difficulties, one participant due to technical issues with video calling, and one due to distress during the first learning trial. Most participants (35/40, 88%) completed all four verbal memory tasks, with two participants completing only two of the tasks due to work or childcare commitments. Overall, data completeness rate was 90%. Three participants reported finding the task difficult, two reported feeling distracted during the encoding tasks, two reported finding the ‘alive or not’ encoding task distracting rather than helpful, and two reported finding that usual memory techniques such as the story technique or keeping words in an auditory loop were difficult to do in this task. One participant commented that they already write a brief diary of things that happened that day.

#### 3.2.6 Online cognitive tests

Few participants completed all 21 cognitive tasks: 6 (15%) participants completed all three cognitive tasks on all 21 occasions. 7 (17.5%) provided complete data for CRT tasks, 8/40 (20%) for the SOS, and 9 (22.5%) for the digit span task. However, on average data completeness was good with 79.8% of allocated online cognitive tasks completed. One participant with PD attempted the tasks but stopped due to frustration at not being able to do the tasks, as they required speed and accuracy. Several participants reported misunderstanding the CRT task and clicking outside of the response window.

#### 3.2.7 Saliva samples

##### 3.2.7.1 Dim-light melatonin assay: Passive drool samples

Most saliva samples (272/280, 97%) were collected as requested. Data completeness rate for analysable samples was 97.1% across the cohort. One participant provided no saliva samples, and 1 participant provided 9/10 samples. Participant-reported sample times were available for 236/272 (86.7%) samples. 120/236 (51%) samples were reported to have been completed within 15 minutes of the scheduled time, as calculated by average bedtimes reported in the sleep diary. On average, participants completed their samples around 7 minutes early (**Table 3**). Previous studies have identified highly variable salivary melatonin secretion, with peaks from 2 pg/mL to 84 pg/mL^57^. Across the cohort, values ranged from beneath detectable levels (<1.37 pg/mL) to 92.4 pg/mL.

**Table 3.**
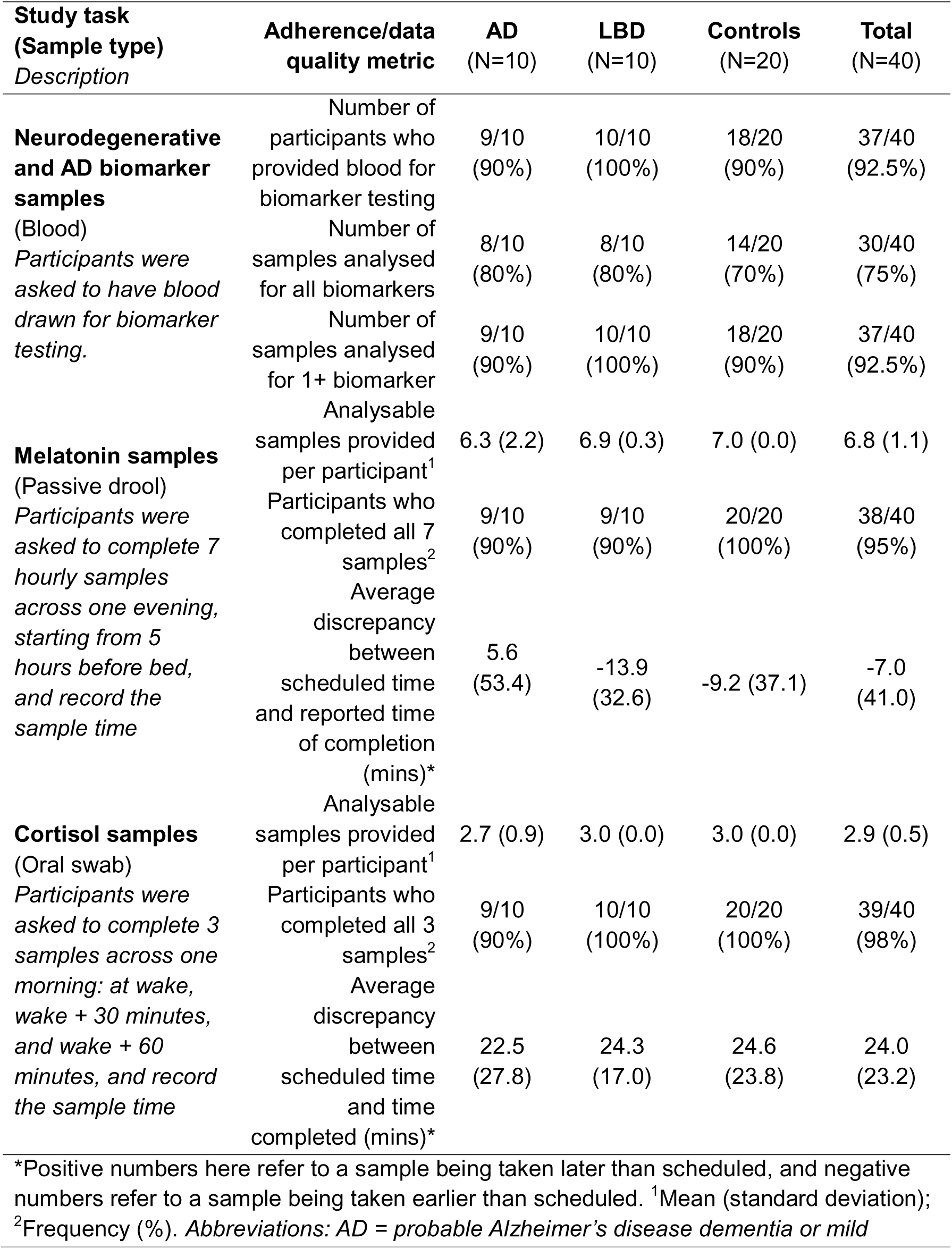
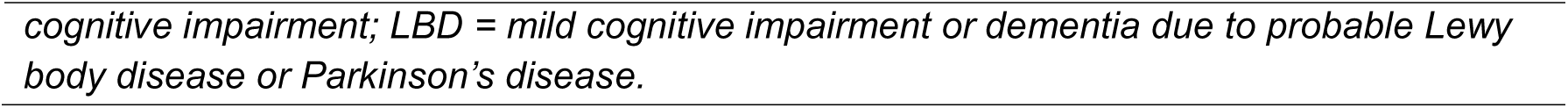
Outcomes relating to data quality and adherence for biomarker analysis: blood biomarkers, salivary dim-light melatonin and salivary cortisol awakening response.

| **Study task**<br>**(Sample type)**<br>*Description* | **Adherence/data**<br>**quality metric** | **AD**<br>**(N=10)** | **LBD**<br>**(N=10)** | **Controls**<br>**(N=20)** | **Total**<br>**(N=40)** |
| --- | --- | --- | --- | --- | --- |
| **Neurodegenerative**<br>**and AD biomarker**<br>**samples**<br>(Blood)<br>*Participants were*<br>*asked to have blood*<br>*drawn for biomarker*<br>*testing.* | Number of<br>participants who<br>provided blood for<br>biomarker testing | 9/10<br>(90%) | 10/10<br>(100%) | 18/20<br>(90%) | 37/40<br>(92.5%) |
|  | Number of<br>samples analysed<br>for all biomarkers | 8/10<br>(80%) | 8/10<br>(80%) | 14/20<br>(70%) | 30/40<br>(75%) |
|  | Number of<br>samples analysed<br>for 1+ biomarker | 9/10<br>(90%) | 10/10<br>(100%) | 18/20<br>(90%) | 37/40<br>(92.5%) |
| **Melatonin samples**<br>(Passive drool)<br>*Participants were*<br>*asked to complete 7*<br>*hourly samples*<br>*across one evening,*<br>*starting from 5*<br>*hours before bed,*<br>*and record the*<br>*sample time* | Analysable<br>samples provided<br>per participant<sup>1</sup> | 6.3 (2.2) | 6.9 (0.3) | 7.0 (0.0) | 6.8 (1.1) |
|  | Participants who<br>completed all 7<br>samples<sup>2</sup> | 9/10<br>(90%) | 9/10<br>(90%) | 20/20<br>(100%) | 38/40<br>(95%) |
|  | Average<br>discrepancy<br>between<br>scheduled time<br>and reported time<br>of completion<br>(mins)* | 5.6<br>(53.4) | -13.9<br>(32.6) | -9.2 (37.1) | -7.0<br>(41.0) |
| **Cortisol samples**<br>(Oral swab)<br>*Participants were*<br>*asked to complete 3*<br>*samples across one*<br>*morning: at wake,*<br>*wake + 30 minutes,*<br>*and wake + 60*<br>*minutes, and record*<br>*the sample time* | Analysable<br>samples provided<br>per participant<sup>1</sup> | 2.7 (0.9) | 3.0 (0.0) | 3.0 (0.0) | 2.9 (0.5) |
|  | Participants who<br>completed all 3<br>samples<sup>2</sup> | 9/10<br>(90%) | 10/10<br>(100%) | 20/20<br>(100%) | 39/40<br>(98%) |
|  | Average<br>discrepancy<br>between<br>scheduled time<br>and time<br>completed (mins)* | 22.5<br>(27.8) | 24.3<br>(17.0) | 24.6<br>(23.8) | 24.0<br>(23.2) |

\*Positive numbers here refer to a sample being taken later than scheduled, and negative numbers refer to a sample being taken earlier than scheduled. <sup>1</sup>Mean (standard deviation);
<sup>2</sup>Frequency (%). Abbreviations: AD = probable Alzheimer's disease dementia or mild
*cognitive impairment; LBD = mild cognitive impairment or dementia due to probable Lewy body disease or Parkinson's disease.*

##### 3.2.7.2 Cortisol awakening response: oral saliva swabs

Participants were instructed to provide saliva samples at wake, after 30 minutes, and after 60 minutes on one morning during the intensive week. Data completeness rate for analysable samples was 96.7% across the cohort. We compared reported saliva sample timings to their scheduled timings, based on final awakening time estimated by EEG. Participant-reported sample times and EEG awakening time was available for 85/117 (72.6%) cortisol samples. Two participants’ EEG recordings (6 samples) suggested wake times later than the first sample recording time and were removed from analysis due to suspected error in recorded saliva timing or date. Participants recorded their first cortisol saliva swab an average of 23 minutes after awakening and overall cortisol samples were generally 24 minutes later than scheduled across the three timepoints (**Table 3**). Previous studies have identified variable salivary cortisol awakening values from 3 µg/L to 19 µg/L in healthy adults^58^. Across the cohort, values ranged from 0.9 µg/L to 13.4 µg/L.

### 3.3 Associations between participant characteristics and adherence

There was some evidence of a weak correlation between MoCA score at baseline and EEG record quality, where those with greater cognitive impairment had lower average EEG record quality (r = 0.31, p = 0.05). Spearman’s rank correlations revealed no evidence of correlations between EEG record quality and age (r = -0.10, p = 0.54), apathy (r = 0.16, p = 0.31), or PSQI score (r= 0.19, p = 0.24). There was also no evidence of correlation between the number of sleep diaries completed and age (r = -0.15, p = 0.35), MoCA score (r = 0.11, p = 0.48), apathy (r = 0.07, p = 0.69), or PSQI score (r = 0.12, p = 0.47).

### 3.4 Resource use

#### 3.4.1 Support from partners and relatives

36% (4/11) of individuals with AD, 85% (11/13) of individuals with PD or LBD, and 10% (2/20) controls attended their consent visit with a partner or relative. 32/40 (80%) participants reported that there would be someone external to the research team (e.g., partner or relative) who could support them with study tasks if needed at baseline. 12/40 (30%) participants reported receiving support with completing study tasks (3 AD, 8 LBD, 1 control). Support from outside of the study team was predominantly from partners and included reminders to complete tasks, device set-up, and troubleshooting technical problems. The participant who withdrew following baseline reported having someone who could support them at home.

#### 3.4.2 Support and contact with the research team

Participants received in-person and remote training and support from a researcher to complete study tasks at baseline (including downloading and setting up applications and turning on recording devices) and offered a refresher training session prior to the intensive week. Participants were also provided with a written instruction manual, scheduled in-app and email reminders, and ad-hoc support as requested by participants or where the team noticed three or more consecutive days of missing sleep diary data. All participants had at least one in-person visit from a researcher (e.g., to download midpoint actigraphy data or retrieve study equipment) during the study period. Participants were advised to contact the research team if they had questions during the study by their preferred method (from email, phone, video call, or instant messaging on the MyDignio app). Support from the research team during the study was most often provided by e-mail and involved responding to participant queries on initial set up (e.g., downloading the MyDignio app), reminders (e.g., tasks to complete or passwords), and technical issues (e.g., links not arriving for Cognitron). Some participants also requested telephone-based, video-based, or home-based support (e.g., to refresh training on the intensive week study tasks). Most participants did not receive regular reminders from the study team to complete tasks.

#### 3.4.3 Device use

One participant was provided with a study tablet upon request, as they owned a suitable device but wanted to keep study activities and apps separate to their personal devices. All other participants used their own smart devices for study activities (i.e., smartphones, tablets, and personal computers). Most participants utilised more than one device to complete online study tasks due to personal preference or convenience. No devices provided by the study team were lost or damaged during the study.

## 4 DISCUSSION

Here, we show that it is feasible to remotely measure sleep and cognition longitudinally in community-dwelling older adults with MCI and dementia due to AD and LBD and healthy older adults. Eligible participants were interested in, enrolled, and remained in the study, despite being asked to complete a high volume of remote and novel study tasks across an eight-week period alongside continuing their usual routines. Only one participant withdrew from the study, and this was before remote data collection started. Most participants were receptive to multimodal home-based research using technology and alternatives to in-lab sleep assessment. Across the cohort, data completeness rate was high and ranged from 79.8% to 97.1%. Our findings support the use of remote, technology-supported research methods to study naturalistic sleep in future trials and indicate some areas where further improvements and refinements would be helpful. With just under half of our sample having possible undiagnosed sleep apnoea, our results also highlight the importance of sleep apnoea screening in older adults, as a risk factor and potentially reversible contributor to cognitive impairment and poor sleep quality^59^.

Many older adults with MCI and dementia routinely leverage technology for cognitive stimulation, performing activities of daily living (such as shopping and banking), entertainment, and socialising and utilise assistive technology solutions such as reminders and navigation to support independence^60^. However, cognitive impairment might impact ability to understand or remember to complete remote or technology-based study tasks, resulting in missing data, and may trigger anxiety if patients feel unable to perform tasks correctly^61^,. Although a few previous studies had examined use of sleep wearables in adults with AD, this was typically done over a brief period, examined feasibility of using only one digital health technology, or required input from a carer or study partner^30-33,62^. Our study demonstrated that older adults with MCI and dementia can successfully complete novel multimodal sleep and cognitive assessments including longitudinal concurrent use of wrist actigraphy, online cognitive tests, digital sleep diaries, and wireless EEG headbands. However, more effort will be required to recruit samples which are representative of the older adult population and those with MCI/dementia, as our study predominantly recruited White males who were familiar with smart technology, albeit not the devices used in RESTED.

Data completeness rates, reflecting both ability to complete study tasks and produce data of analysable quality, were high across all study tasks and participants. Remote data collection comes with a risk of missing data or non-adherence, however we were able to mitigate this risk by using devices which upload data to servers at regular intervals (e.g., Dreem, Dignio) and, less conveniently, by downloading data manually from devices to check compliance and data quality and offer support where needed (e.g., actigraphy). Passive monitoring devices, as well as devices which upload to servers automatically, simultaneously reduce burden on participants and researchers whilst minimising data loss by allowing researchers to monitor and respond quickly to any user or technical issues.

Reasons for missing data were usually known and typically related to infrequent but intentional decisions to remove a device (e.g., due to a social event or illness) or, in most cases, technical issues with devices or software beyond the participants’ control, which would not introduce bias. Although we observed a weak correlation between EEG record quality and baseline cognitive impairment, we observed these issues across all participant subgroups and overall record quality remained high.

Use of remote sleep and cognitive monitoring can help to partly or fully de-centralise research, as participants can utilise their own devices or be provided with technology via post where needed^63^ and are more scalable for clinical trials^64^. Frequent or lengthy clinic visits, particularly those which require overnight stays for sleep analysis, can be burdensome for participants and may reduce inclusivity by requiring participants to live near a study site or travel long distances. More frequent but briefer remote assessments may also help to mitigate against the risk of large amounts of missing data resulting from a missed clinic appointment, can act as “digital biomarkers” that may be able to detect changes more sensitively than an annual follow-up^65^ and are more convenient for participants^66^. Though not the focus of our work, research could also explore the use of digital tools to assess fluctuations in cognition over the day in LBD (where fluctuation in cognition is a key diagnostic criteria) and AD (where a sundowning effect of heightened distress is often observed during the evening hours) – both of which are difficult to assess in clinical settings ^67,68^.

It has been argued that study partners are essential for participant safety and wellbeing in dementia research, even at the preclinical stage where cognitively normal individuals are enrolled^69^. However, most participants provided good quality data with minimal to no input from others, and several participants would not have been able to participate if there was a study partner requirement. Our findings promote inclusive research designs with an option to formally recruit and collect data from study partners, for support and information to corroborate outcomes from patients if required.

There are an increasing number of options when considering how to measure sleep and cognition from home including both research-grade and consumer sleep trackers^70 71^. We opted to utilise existing technologies which were non-invasive, affordable, required minimal training or supervision, were commercially available research-grade or consumer-grade devices which met necessary data privacy regulations, and provided data in a format which could be analysed using open-source software. Existing technologies can be implemented immediately, reducing time and cost to set up research studies, often have a more mature user interface and are less error-prone than a newly developed solution, and crucially may be easier to compare amongst studies for meta-analysis. Several study tasks required ongoing technical support or services from the manufacturers and/or developers. As a customer rather than a collaborator, we could not always identify or respond to technical problems and with some providers we were not informed in advance of several significant changes which impacted the study or its participants resulting in data loss. Collaborating with industry, ideally at an early stage of the research process, might ensure longevity and continued support throughout the life course of the study and increased flexibility to adapt technology to better fit research (e.g., data validation for digital patient-reported outcomes).

### 4.1 Limitations

Women, older adults (80+), and minoritised ethnic groups were under-represented in our study, which is often observed in dementia research^72^. Though some barriers (e.g., mistrust and accessibility of research) and motivators (e.g., altruism) have been identified, further work is needed to identify how to improve research access, inclusion, and participation in dementia research^73^. There may also have been a self-selection bias, whereby individuals who were likely to be more competent and enthusiastic about using technology volunteered to participate in the study. Most of our participants were regular smartphone users, and several prospective participants declined to participate due to concerns regarding their confidence or interest in using the technologies. Though the proportion of older adults in the UK using the internet is increasing, a significant proportion of older adults do not regularly use the internet or lack fundamental skills such as being able to turn on a device and enter login details^34^. Older adults with MCI and dementia who regularly use devices report that smartphones and tablets can be useful to support activities of daily living, as well as for communication, entertainment, and recreation, but also list concerns including cybersecurity and vulnerability to fraud^74^. Education around sleep and brain health and basic digital skills training on how devices can support individuals living with cognitive impairment may improve perceptions of capability and motivation to participate in similar studies^56,74^. Exploring the feasibility of more passive monitoring technologies, such as mattress sensors or smartphone-based passive sensing which require less technical knowledge or engagement may help to increase participation and confidence from those with less experience or interest in technology, though these techniques may invite additional concerns around data privacy^75^. Increasing the pool of prospective participants (e.g., recruiting via multiple sites and meaningful engagement with community groups) and considering digital inclusivity is likely to enhance recruitment in future studies. Finally, we did not set *a priori* feasibility cut-offs for study tasks. Despite these limitations, the high completion and retention rate from the RESTED study suggests that remote sleep and cognitive monitoring could offer detailed sleep profiling suitable for tracking change in sleep over disease progression or for monitoring change in sleep clinical trials in older adults with or at risk of dementia.

### 4.2 Conclusion

We conclude that it is feasible for older adults with MCI and dementia, as well as healthy older adults, to engage in multimodal remote sleep and cognitive research including using wireless EEG, actigraphy, and mobile or web applications. Remote research methods offer the opportunity to study naturalistic sleep and its relationship with cognition and dementia over extended periods of time, are scalable, and should be considered when designing future clinical trials in sleep and dementia.

## Data Availability

All data produced in the present study are available upon reasonable request to the authors.

## 6 ACKNOWLEDGEMENTS, SOURCES OF FUNDING, AND DISCLOSURES

### 6.1 Acknowledgements

The authors sincerely thank the RESTED study participants and their relatives and partners who supported them to take part. We would also like to thank the ReMemBr group Lived Experience Experts for their contributions to improving the study design and supporting us to make the study as accessible as possible. We would also like to thank the North Bristol NHS Trust Respiratory Physiology department. Finally, we would like to thank the teams at Dreem Research, Cognitron, Join Dementia Research, and Ewa G Truchanowicz and Phil Reay and their teams at Dignio UK.

### 6.2 Sources of funding

The RESTED study was funded by charitable organisations (BRACE, Alzheimer’s Research UK, the Bristol & Weston Hospitals Charity (previously Above & Beyond), and the David Telling Charitable Trust), the NIHR Bristol Biomedical Research Centre, and philanthropic donations from S. Scobie and A. Graham. JB also received funding from Alzheimer’s Research UK (ARUK) supported by the Margaret Jost Fellowship and the Don Thoburn Memorial Scholarship. The funders had no role in the trial design, patient recruitment, data collection, analysis, interpretation, nor in writing of the manuscript or the decision to submit it for publication.

### 6.3 Conflict of interest

The authors declare no conflicts of interest. EC has received funding from Biogen, Eisai, and Lilly for consultancy and providing educational resources.

### 6.4 Consent statement

All participants provided written informed consent prior to participating in the study.

## Notes

### Author Declarations

The Research Ethics Committee of Yorkshire and the Humber - Bradford Leeds gave ethical approval for this work on behalf of the Health Research Authority (reference 21/YH/0177).

## 5 REFERENCES

1. Koren T, Fisher E, Webster L, Livingston G, Rapaport P. Prevalence of sleep disturbances in people with dementia living in the community: A systematic review and meta-analysis. Ageing Research Reviews. 2023;83:101782. doi: 10.1016/j.arr.2022.101782

2. Postuma RB, Iranzo A, Hu M, et al. Risk and predictors of dementia and parkinsonism in idiopathic REM sleep behaviour disorder: a multicentre study. Brain. 2019;142(3):744–759. doi:10.1093/brain/awz030

3. Harenbrock J, Holling H, Reid G, Koychev I. A meta-analysis of the relationship between sleep and β-Amyloid biomarkers in Alzheimer’s disease. Biomarkers in Neuropsychiatry. 2023;9:100068. doi: 10.1016/j.bionps.2023.100068

4. Zamore Z, Veasey SC. Neural consequences of chronic sleep disruption. Trends Neurosci. 2022;45(9):678–691. doi:10.1016/j.tins.2022.05.007

5. Zielinski MR, Gibbons AJ. Neuroinflammation, Sleep, and Circadian Rhythms. Front Cell Infect Microbiol. 2022;12:853096. doi:10.3389/fcimb.2022.853096

6. Eshera YM, Gavrilova L, Hughes JW. Sleep is Essential for Cardiovascular Health: An Analytic Review of the Relationship Between Sleep and Cardiovascular Mortality. Am J Lifestyle Med. 2024;18(3):340–350. doi:10.1177/15598276231211846

7. Lee VV, Schembri R, Jordan AS, Jackson ML. The independent effects of sleep deprivation and sleep fragmentation on processing of emotional information. Behavioural Brain Research. 2022;424:113802. doi: 10.1016/j.bbr.2022.113802

8. Dzierzewski JM, Dautovich N, Ravyts S. Sleep and Cognition in Older Adults. Sleep Med Clin. Mar 2018;13(1):93–106. doi:10.1016/j.jsmc.2017.09.009

9. Seda G, Matwiyoff G, Parrish JS. Effects of obstructive sleep apnea and CPAP on cognitive function. Current Neurology and Neuroscience Reports. 2021;21(7):32.

10. Costa YS, Lim ASP, Thorpe KE, et al. Investigating changes in cognition associated with the use of CPAP in cognitive impairment and dementia: A retrospective study. Sleep Medicine. 2023;101:437–444. doi: 10.1016/j.sleep.2022.11.037

11. Liu WT, Huang HT, Hung HY, et al. Continuous Positive Airway Pressure Reduces Plasma Neurochemical Levels in Patients with OSA: A Pilot Study. Life (Basel). 2023;13(3)doi:10.3390/life13030613

12. Sabia S, Fayosse A, Dumurgier J, et al. Association of sleep duration in middle and old age with incidence of dementia. Nat Commun. 2021;12(1):2289. doi:10.1038/s41467-021-22354-2

13. Mayer G, Frohnhofen H, Jokisch M, Hermann DM, Gronewold J. Associations of sleep disorders with all-cause MCI/dementia and different types of dementia - clinical evidence, potential pathomechanisms and treatment options: A narrative review. Front Neurosci. 2024;18:1372326. doi:10.3389/fnins.2024.1372326

14. Clynes J, Blackman J, Swirski M, Leng Y, Harding S, Coulthard E. Interventions to enhance sleep in mild cognitive impairment and mild Alzheimer’s dementia: a systematic review. Sleep medicine. 2019;64:-S77. doi:10.1016/j.sleep.2019.11.212

15. Blackman J, Morrison HD, Lloyd K, et al. The past, present, and future of sleep measurement in mild cognitive impairment and early dementia—Towards a core outcome set: A scoping review. Sleep: Journal of Sleep and Sleep Disorders Research. 45(7):1–11. doi:10.1093/sleep/zsac077

16. Kainec KA, Caccavaro J, Barnes M, Hoff C, Berlin A, Spencer RMC. Evaluating Accuracy in Five Commercial Sleep-Tracking Devices Compared to Research-Grade Actigraphy and Polysomnography. Sensors (Basel). 2024;24(2)doi:10.3390/s24020635

17. Tadokoro K, Ohta Y, Hishikawa N, et al. Discrepancy of subjective and objective sleep problems in Alzheimer’s disease and mild cognitive impairment detected by a home-based sleep analysis. Journal of Clinical Neuroscience. 2020;74:76–80. doi: 10.1016/j.jocn.2020.01.085

18. Casagrande M, Forte G, Favieri F, Corbo I. Sleep Quality and Aging: A Systematic Review on Healthy Older People, Mild Cognitive Impairment and Alzheimer’s Disease. International Journal of Environmental Research and Public Health. 2022;19(14):8457.

19. Williams JM, Kay DB, Rowe M, McCrae CS. Sleep Discrepancy, Sleep Complaint, and Poor Sleep Among Older Adults. The Journals of Gerontology: Series B. 2013;68(5):712–720. doi:10.1093/geronb/gbt030

20. Markun LC, Sampat A. Clinician-Focused Overview and Developments in Polysomnography. Current Sleep Medicine Reports. 2020;6(4):309–321. doi:10.1007/s40675-020-00197-5

21. Goparaju B, de Palma G, Bianchi MT. Naturalistic sleep tracking in a longitudinal cohort: how long is long enough? medRxiv. 2024:2024.10.19.24315818. doi:10.1101/2024.10.19.24315818

22. Blackman J, Morrison HD, Lloyd K, et al. Sleep Measurement Heterogeneity in Mild Cognitive Impairment and Early Dementia - Towards a Core Outcome Set: A Scoping Review. Sleep medicine. 2022;100:S148–S149. doi:10.1016/j.sleep.2022.05.403

23. Green SF, Frame T, Banerjee LV, et al. A systematic review of the validity of non-invasive sleep-measuring devices in mid-to-late life adults: Future utility for Alzheimer’s disease research. Sleep Medicine Reviews. 2022;65:101665. doi: 10.1016/j.smrv.2022.101665

24. Kourtis LC, Regele OB, Wright JM, Jones GB. Digital biomarkers for Alzheimer’s disease: the mobile/wearable devices opportunity. npj Digital Medicine. 2019/02/21 2019;2(1):9. doi:10.1038/s41746-019-0084-2

25. Arnal PJ, Thorey V, Debellemaniere E, et al. The Dreem Headband compared to polysomnography for electroencephalographic signal acquisition and sleep staging. Sleep. Nov 12 2020;43(11). doi:10.1093/sleep/zsaa097

26. Miller DJ, Sargent C, Roach GD. A Validation of Six Wearable Devices for Estimating Sleep, Heart Rate and Heart Rate Variability in Healthy Adults. Sensors. 2022;22(16):6317.

27. Cay G, Ravichandran V, Sadhu S, et al. Recent Advancement in Sleep Technologies: A Literature Review on Clinical Standards, Sensors, Apps, and AI Methods. IEEE Access. 2022;10:104737–104756. doi:10.1109/ACCESS.2022.3210518

28. Ding Z, Lee TL, Chan AS. Digital Cognitive Biomarker for Mild Cognitive Impairments and Dementia: A Systematic Review. J Clin Med. 2022;11(14)doi:10.3390/jcm11144191

29. Masanneck L, Gieseler P, Gordon WJ, Meuth SG, Stern AD. Evidence from ClinicalTrials.gov on the growth of Digital Health Technologies in neurology trials. npj Digital Medicine. 2023;6(1):23. doi:10.1038/s41746-023-00767-1

30. Kent BA, Casciola AA, Carlucci SK, et al. Home EEG sleep assessment shows reduced slow-wave sleep in mild–moderate Alzheimer’s disease. Alzheimer’s & Dementia: Translational Research & Clinical Interventions. 2022;8(1):e12347. doi: 10.1002/trc2.12347

31. Pavlickova H, Russell AE, Lightman S, McCabe R. Feasibility of salivary cortisol collection in patients and companions attending dementia diagnostic meetings in memory clinics. BMC Research Notes. 2021/01/21 2021;14(1):30. doi:10.1186/s13104-021-05446-6

32. Jones C, Moyle W. A feasibility study of Dreampad™ on sleep, wandering and agitated behaviors in people living with dementia. Geriatric Nursing. 2020;41(6):782–789. doi:10.1016/j.gerinurse.2020.04.014

33. Guu T-W, Brem A-K, Albertyn CP, Kandangwa P, Aarsland D, ffytche D. Wrist-worn actigraphy in agitated late-stage dementia patients: A feasibility study on digital inclusion. Alzheimer’s & Dementia. 2024;20(5):3211–3218. doi:10.1002/alz.13772

34. UK A. Facts and figures about digital inclusion and older people. 2024. June 2024. Accessed 16/01/2025. https://www.ageuk.org.uk/siteassets/documents/reports-and-publications/reports-and-briefings/active-communities/internet-use-statistics-june-2024.pdf

35. Holthe T, Halvorsrud L, Karterud D, Hoel K-A, Lund A. Usability and acceptability of technology for community-dwelling older adults with mild cognitive impairment and dementia: a systematic literature review. Clinical interventions in aging. 2018;13:863–886. doi:10.2147/CIA.S154717

36. Chien S-Y, Zaslavsky O, Berridge C. Technology Usability for People Living With Dementia: Concept Analysis. JMIR Aging. 2024;7:e51987. doi:10.2196/51987

37. Gabb VG, Blackman J, Morrison HD, et al. Remote Evaluation of Sleep and Circadian Rhythms in Older Adults With Mild Cognitive Impairment and Dementia: Protocol for a Feasibility and Acceptability Mixed Methods Study. JMIR research protocols. 2024;13:e52652. doi: 10.2196/52652

38. McKhann GM, Knopman DS, Chertkow H, et al. The diagnosis of dementia due to Alzheimer’s disease: recommendations from the National Institute on Aging-Alzheimer’s Association workgroups on diagnostic guidelines for Alzheimer’s disease. Alzheimers Dement. 2011;7(3):263–9. doi:10.1016/j.jalz.2011.03.005

39. Albert MS, DeKosky ST, Dickson D, et al. The diagnosis of mild cognitive impairment due to Alzheimer’s disease: Recommendations from the National Institute on Aging-Alzheimer’s Association workgroups on diagnostic guidelines for Alzheimer’s disease. Alzheimer’s & Dementia. 2011;7(3):270–279. doi:10.1016/j.jalz.2011.03.008

40. Yamada M, Komatsu J, Nakamura K, et al. Diagnostic Criteria for Dementia with Lewy Bodies: Updates and Future Directions. J Mov Disord. 2020;13(1):1–10. doi:10.14802/jmd.19052

41. Litvan I, Goldman JG, Tröster AI, et al. Diagnostic criteria for mild cognitive impairment in Parkinson’s disease: Movement Disorder Society Task Force guidelines. Mov Disord. 2012;27(3):349–56. doi:10.1002/mds.24893

42. Blackman J, Morrison HD, Gabb V, et al. Remote evaluation of sleep to enhance understanding of early dementia due to Alzheimer’s Disease (RESTED-AD): an observational cohort study protocol. BMC Geriatr. 2023;23(1):590. doi:10.1186/s12877-023-04288-0

43. Blackman J, Swirski M, Clynes J, Harding S, Leng Y, Coulthard E. Pharmacological and non-pharmacological interventions to enhance sleep in mild cognitive impairment and mild Alzheimer’s disease: A systematic review. Journal of sleep research. 30(4):20. doi:10.1111/jsr.13229

44. Blackman J, Gabb V, Carrigan N, et al. Sleep quality during and after severe acute respiratory syndrome coronavirus 2 (COVID-19) lockdowns in the UK: Results from the SleepQuest study. Journal of sleep research. 2024;33:e14205. doi:10.1111/jsr.14205

45. Buysse DJ, Reynolds CF, 3rd, Monk TH, Berman SR, Kupfer DJ. The Pittsburgh Sleep Quality Index: a new instrument for psychiatric practice and research. Psychiatry Res. 1989;28(2):193–213. doi:10.1016/0165-1781(89)90047-4

46. Johns MW. A new method for measuring daytime sleepiness: The Epworth Sleepiness Scale. Sleep: Journal of Sleep Research & Sleep Medicine. 1991;14(6):540–545. doi:10.1093/sleep/14.6.540

47. Chung F, Abdullah HR, Liao P. STOP-Bang Questionnaire: A Practical Approach to Screen for Obstructive Sleep Apnea. Chest. 2016;149(3):631–8. doi:10.1378/chest.15-0903

48. Sheikh J, Yesavage J. Geriatric Depression Scale (GDS): recent findings and development of a shorter version In: Brink T, ed. Clinical Gerontology: A Guide to Assessment and Intervention. New York: Howarth Press; 1986.

49. Spitzer RL, Kroenke K, Williams JB, Löwe B. A brief measure for assessing generalized anxiety disorder: the GAD-7. Arch Intern Med. May 22 2006;166(10):1092–7. doi:10.1001/archinte.166.10.1092

50. Marin RS, Biedrzycki RC, Firinciogullari S. Reliability and validity of the apathy evaluation scale. Psychiatry Research. 1991;38(2):143–162. doi:10.1016/0165-1781(91)90040-V

51. Carney CE, Buysse DJ, Ancoli-Israel S, et al. The consensus sleep diary: standardizing prospective sleep self-monitoring. Sleep. Feb 1 2012;35(2):287–302. doi:10.5665/sleep.1642

52. Ravindran KKG, della Monica C, Atzori G, et al. Evaluation of Dreem headband for sleep staging and EEG spectral analysis in people living with Alzheimer’s and older adults. medRxiv. 2024:2024.12.18.24319240. doi:10.1101/2024.12.18.24319240

53. González DA, Wang D, Pollet E, et al. Performance of the Dreem 2 EEG headband, relative to polysomnography, for assessing sleep in Parkinson’s disease. Sleep Health. 2024;10(1):24–30. doi:10.1016/j.sleh.2023.11.012

54. Migueles JH, Rowlands AV, Huber F, Sabia S, van Hees VT. GGIR: A Research Community–Driven Open Source R Package for Generating Physical Activity and Sleep Outcomes From Multi-Day Raw Accelerometer Data. Journal for the Measurement of Physical Behaviour. 2019 2019;2(3):188–196. doi:10.1123/jmpb.2018-0063

55. van Hees VT, Sabia S, Jones SE, et al. Estimating sleep parameters using an accelerometer without sleep diary. Scientific Reports. 2018/08/28 2018;8(1):12975. doi:10.1038/s41598-018-31266-z

56. Michie S, van Stralen MM, West R. The behaviour change wheel: A new method for characterising and designing behaviour change interventions. Implementation Science. 2011;6(1):42. doi:10.1186/1748-5908-6-42

57. Burgess HJ, Fogg LF. Individual Differences in the Amount and Timing of Salivary Melatonin Secretion. PLOS ONE. 2008;3(8):e3055. doi:10.1371/journal.pone.0003055

58. Trilck M, Flitsch J, Lüdecke DK, Jung R, Petersenn S. Salivary Cortisol Measurement - a Reliable Method for the Diagnosis of Cushing’s Syndrome. Exp Clin Endocrinol Diabetes. 2005;113(04):225–230. doi:10.1055/s-2005-837667

59. Naismith S, Winter V, Gotsopoulos H, Hickie I, Cistulli P. Neurobehavioral Functioning in Obstructive Sleep Apnea: Differential Effects of Sleep Quality, Hypoxemia and Subjective Sleepiness. Journal of Clinical and Experimental Neuropsychology. 2004;26(1):43–54. doi:10.1076/jcen.26.1.43.23929

60. Holthe T, Halvorsrud L, Lund A. Digital Assistive Technology to Support Everyday Living in Community-Dwelling Older Adults with Mild Cognitive Impairment and Dementia. Clin Interv Aging. 2022;17:519–544. doi:10.2147/cia.S357860

61. Ahuja M, Siddhpuria S, Reppas-Rindlisbacher C, et al. Sleep monitoring challenges in patients with neurocognitive disorders: A cross-sectional analysis of missing data from activity trackers. Health Sci Rep. 2022;5(3):e608. doi:10.1002/hsr2.608

62. Van Den Berg JF, Van Rooij FJ, Vos H, et al. Disagreement between subjective and actigraphic measures of sleep duration in a population-based study of elderly persons. J Sleep Res. 2008;17(3):295–302. doi:10.1111/j.1365-2869.2008.00638.x

63. Aiyegbusi OL, Davies EH, Myles P, et al. Digitally enabled decentralised research: opportunities to improve the efficiency of clinical trials and observational studies. BMJ Evidence-Based Medicine. 2023;28(5):328–331. doi:10.1136/bmjebm-2023-112253

64. Inan OT, Tenaerts P, Prindiville SA, et al. Digitizing clinical trials. npj Digital Medicine. 2020;3(1):101. doi:10.1038/s41746-020-0302-y

65. Dodge HH, Zhu J, Mattek NC, Austin D, Kornfeld J, Kaye JA. Use of High-Frequency In-Home Monitoring Data May Reduce Sample Sizes Needed in Clinical Trials. PLoS One. 2015;10(9):e0138095. doi:10.1371/journal.pone.0138095

66. Black BS, Taylor HA, Rabins PV, Karlawish J. Study partners perform essential tasks in dementia research and can experience burdens and benefits in this role. Dementia (London). May 13 2016; doi:10.1177/1471301216648796

67. O’Dowd S, Schumacher J, Burn DJ, et al. Fluctuating cognition in the Lewy body dementias. Brain. 2019;142(11):3338–3350. doi:10.1093/brain/awz235

68. Canevelli M, Valletta M, Trebbastoni A, et al. Sundowning in Dementia: Clinical Relevance, Pathophysiological Determinants, and Therapeutic Approaches. Mini Review. Frontiers in Medicine. 2016;3doi:10.3389/fmed.2016.00073

69. Grill JD, Karlawish J. Study partners should be required in preclinical Alzheimer’s disease trials. Alzheimers Res Ther. 2017;9(1):93. doi:10.1186/s13195-017-0327-x

70. de Zambotti M, Goldstein C, Cook J, et al. State of the science and recommendations for using wearable technology in sleep and circadian research. Sleep. 2023;47(4)doi:10.1093/sleep/zsad325

71. Lee T, Cho Y, Cha KS, et al. Accuracy of 11 Wearable, Nearable, and Airable Consumer Sleep Trackers: Prospective Multicenter Validation Study. JMIR Mhealth Uhealth. 2023;11:e50983. doi:10.2196/50983

72. Banzi R, Camaioni P, Tettamanti M, Bertele’ V, Lucca U. Older patients are still under-represented in clinical trials of Alzheimer’s disease. Alzheimer’s Research & Therapy. 2016;8(1):32. doi:10.1186/s13195-016-0201-2

73. Gilmore-Bykovskyi AL, Jin Y, Gleason C, et al. Recruitment and retention of underrepresented populations in Alzheimer’s disease research: A systematic review. Alzheimers Dement (N Y). 2019;5:751–770. doi:10.1016/j.trci.2019.09.018

74. Wilson SA, Byrne P, Rodgers SE. ‘I’d be lost without my smartphone’: a qualitative analysis of the use of smartphones and tablets by people living with dementia, mild cognitive impairment, and their caregivers. Aging & Mental Health. 2024;28(4):595–603. doi:10.1080/13607863.2023.2205585

75. Cornet VP, Holden RJ. Systematic review of smartphone-based passive sensing for health and wellbeing. J Biomed Inform. 2018;77:120–132. doi:10.1016/j.jbi.2017.12.008

