## Supplementary Files for "Longitudinal remote sleep and cognitive research in older adults with mild cognitive impairment and dementia: a prospective feasibility cohort study"

### Supplementary Materials 1

STROBE Statement—checklist of items that should be included in reports of observational studies

|  | Item No | Recommendation | Page  No |
| --- | --- | --- | --- |
| **Title and abstract** | 1 | (*a*) Indicate the study’s design with a commonly used term in the title or the abstract | 1 |
|  |  | (*b*) Provide in the abstract an informative and balanced summary of what was done and what was found | 3 |
| Introduction | | | |
| Background/rationale | 2 | Explain the scientific background and rationale for the investigation being reported | 4-5 |
| Objectives | 3 | State specific objectives, including any prespecified hypotheses | 6 |
| Methods | | | |
| Study design | 4 | Present key elements of study design early in the paper | 6 |
| Setting | 5 | Describe the setting, locations, and relevant dates, including periods of recruitment, exposure, follow-up, and data collection | 7 |
| Participants | 6 | (*a*) Give the eligibility criteria, and the sources and methods of selection of participants. Describe methods of follow-up | 7-8 |
| Variables | 7 | Clearly define all outcomes, exposures, predictors, potential confounders, and effect modifiers. Give diagnostic criteria, if applicable | 8-12 |
| Data sources/ measurement | 8* | For each variable of interest, give sources of data and details of methods of assessment (measurement). Describe comparability of assessment methods if there is more than one group | 8-12 |
| Bias | 9 | Describe any efforts to address potential sources of bias | 12 |
| Study size | 10 | Explain how the study size was arrived at | 8 |
| Quantitative variables | 11 | Explain how quantitative variables were handled in the analyses. If applicable, describe which groupings were chosen and why | 12 |
| Statistical methods | 12 | (*a*) Describe all statistical methods, including those used to control for confounding | 12 |
|  |  | (*b*) Describe any methods used to examine subgroups and interactions | 12 |
|  |  | (*c*) Explain how missing data were addressed | 12 |
|  |  | (*d*) *Cohort study*—If applicable, explain how loss to follow-up was addressed | n/a |
|  |  | (*e*) Describe any sensitivity analyses | n/a |

| Results | | | |
| --- | --- | --- | --- |
| Participants | 13 | (a) Report numbers of individuals at each stage of study—e.g., numbers potentially eligible, examined for eligibility, confirmed eligible, included in the study, completing follow-up, and analysed | Figure |
|  |  | (b) Give reasons for non-participation at each stage | Figure |
|  |  | (c) Consider use of a flow diagram | Figure |
| Descriptive data | 14* | (a) Give characteristics of study participants (eg demographic, clinical, social) and information on exposures and potential confounders | 14 |
|  |  | (b) Indicate number of participants with missing data for each variable of interest | 17,18 |
|  |  | (c) Summarise follow-up time (e.g., average and total amount) | 13 |
| Outcome data | 15 | Report numbers of outcome events or summary measures over time | 13-28 |
| Main results | 16 | (*a*) Give unadjusted estimates and, if applicable, confounder-adjusted estimates and their precision (eg, 95% confidence interval). Make clear which confounders were adjusted for and why they were included | 13-28 |
|  |  | (*b*) Report category boundaries when continuous variables were categorized | n/a |
|  |  | (*c*) If relevant, consider translating estimates of relative risk into absolute risk for a meaningful time period | n/a |
| Other analyses | 17 | Report other analyses done—e.g., analyses of subgroups and interactions, and sensitivity analyses | 14-28 |
| Discussion | | | |
| Key results | 18 | Summarise key results with reference to study objectives | 21-25 |
| Limitations | 19 | Discuss limitations of the study, taking into account sources of potential bias or imprecision. Discuss both direction and magnitude of any potential bias | 25-26 |
| Interpretation | 20 | Give a cautious overall interpretation of results considering objectives, limitations, multiplicity of analyses, results from similar studies, and other relevant evidence | 26 |
| Generalisability | 21 | Discuss the generalisability (external validity) of the study results | 25 |
| Other information | | | |
| Funding | 22 | Give the source of funding and the role of the funders for the present study and, if applicable, for the original study on which the present article is based | 32 |

### Supplementary Materials 2

Further information is provided on the devices used to measure sleep and circadian rhythms.

| **Home sleep and circadian monitoring study task** | **Device details** | **Schedule for data collection** |
| --- | --- | --- |
| Actigraphy watch | Axivity AX3 with accompanying wristwatch strap | 56 nights, continuous |
| Wireless EEG headband | Dreem 2 dry-electrode EEG headband | 7 nights, intensive week only |
| Saliva swabs (for cortisol) | Salimetrics® Oral Swabs (Cat. 5001.02)  Pre-labelled Swab Storage Tubes (Cat. 5001.05)  Paper diary for timings | 3 samples across 1 morning in the intensive week |
| Passive drool (for melatonin) | SalivaBio Saliva Collection Aids (Cat. 5016.04)  Pre-labelled Cryovial 2mL (Cat. 5004.01)  Paper diary for timings | 7 samples across 1 evening in the intensive week |
| Overnight pulse oximetry | Nonin WristOx 2® Model 3150 pulse oximeter | 2 consecutive nights across the entire study |
| Study tablet (provided if requested) | Apple iPad | n/a |

### Supplementary Materials 3

|  | F7_O1 | F8_O2 | Fp1_F8 | F8_F7 | F8_O1 | F7_O2 | Fp1_F7 | Fp1_O1 | Fp1_O2 |
| --- | --- | --- | --- | --- | --- | --- | --- | --- | --- |
| AD/aMCI | 50.21 | 45.23 | 68.40 | 73.24 | 41.27 | 40.86 | 71.56 | 64.60 | 46.13 |
| LBD | 50.89 | 47.69 | 68.05 | 75.79 | 19.68 | 22.67 | 70.78 | 54.36 | 57.14 |
| Control | 72.31 | 64.48 | 83.77 | 89.67 | NA | NA | 84.18 | 64.74 | 57.44 |
| Data from channels F7_O1, F8_O2,Fp1_F8, F8_F7, and Fp1_F7 were calculated for all participants. Participants enrolled earliest in the recruitment period also had channels for F8_O1 and F7_O2, which had the lowest record quality, whilst for later participants, Dreem switched to providing data for channels Fp1_O1 and Fp1_O2 which had higher record quality. | | | | | | | | | |

### Supplementary Materials 4

Additional information on reasons for missing data and the affected study tasks is provided.

| Reason for missing data | Study task(s) affected | Example |
| --- | --- | --- |
| **Cognitive**  Difficulty following instructions for completing a task  Discontinuing a task due to frustration or distress  Forgetting to complete (or if they had already completed) a task | Wireless EEG headband, actigraphy, cognitive tests, verbal memory tasks  Cognitive tests, verbal memory tasks  Verbal memory task, sleep diary, wireless EEG headband | Participant wore EEG headband but did not follow the instructions to initiate the recording  Participant was not able to focus or remember the task during the word list task presentation so asked to stop  Participant took off the actigraphy watch to bathe and forgot to put it back on |
| **Lifestyle / social**  Planning around usual routines  Perception of others at social events | Saliva samples, verbal memory tasks, cognitive tests  Actigraphy | Participant with childcare responsibilities during the evenings was unable to complete evening memory task calls with a researcher  Participant removed the actigraph as they felt self-conscious to wear it at a formal event |
| **Physiological**  Unable to obtain (sufficient) sample from participant  Unrelated physical illness | Saliva samples, blood draw  Wireless EEG headband, blood draw | Participant was unable to produce sufficient saliva due to dry mouth  Participant skipped a night’s recording due to feeling generally unwell |
| **Physical**  Difficulty physically completing a task  Discomfort triggered device removal | Saliva samples, cognitive tests  Pulse oximetry, actigraphy | Participant could not open the tube for the saliva sample  Participants reported itching on wrist underneath actigraph so took a break from wearing it until itching resolved |
| **Technical**  Battery or device failure  Issues accessing website and/or mobile application due to software upgrades  Issues during sample processing  Technical issues unrelated to the study | Actigraphy, pulse oximetry  Cognitive tests, sleep diaries  Saliva samples, plasma biomarkers  Verbal memory tasks | Actigraphy battery failure caused by long-term storage  Software update caused previous data to be stored in the cache and prohibited new data entry  Melatonin level beneath detectable levels for assay  Participant had an unstable internet connection and was unable to get the volume or microphone on their device to work so could not participate in the verbal memory tasks |
